# An ancestry-enriched HNF4A variant and GP2 reveal distinct mechanisms of type 2 diabetes in exome-wide study of 13,674 cases and 41,024 controls

**DOI:** 10.1101/2025.09.24.25336527

**Authors:** Sam Hodgson, Vi Bui, Siqi Hu, Cyrielle Maroteau, Margherita Bigossi, Alicia Huerta-Chagoya, Trang Nguyen, Adem Y. Dawed, Ryan Koesterer, Maheak Vora, Daniel Stow, Alice Williamson, Alexandra M Blee, Julia Carrasco-Zanini Sanchez, Viswanathan Baskar, Saravanan Jebarani, Benjamin M Jacobs, Georgios Kalantzis, Stuart Rison, Klaudia Walter, Erwan Pennarun, Katherine Taylor, Sarah Hsu, Genes & Health Research Team, MDRF Research Team, Alisa Manning, Miriam Udler, Hilary C Martin, Inês Barroso, Jason Flannick, Matteo Fumagalli, Venkatesan Radha, Rajendra Pradeepa, Claudia Langenberg, Viswanathan Mohan, Ranjit Mohan Anjana, David A van Heel, Josep M. Mercader, Yalda Jamshidi, Sarah Finer, Amit R. Majithia, Moneeza K Siddiqui

**Affiliations:** Wolfson Institute of Population Health, Queen Mary University of London, United Kingdom; Division of Endocrinology & Metabolism, University of San Diego School of Medicine; Novo Nordisk Research Centre Oxford, Oxford, United Kingdom; Broad Institute of Harvard and MIT; Precision Healthcare University Research Institute, Queen Mary University of London, United Kingdom; Berlin Institute of Health at Charite, Universitätsmedizin, Berlin, Germany; Madras Diabetes Research Foundation (ICMR- Collaborating Centre of Excellence), Chennai, India; Wellcome Sanger Institute, Hinxton, United Kingdom; Exeter Centre of Excellence for Diabetes Research (EXCEED), University of Exeter Medical School, Exeter, United Kingdom; School of Biological and Behavioural Sciences, Queen Mary University of London, United Kingdom; Dr. Mohan’s Diabetes Specialities Centre (IDF Centre of Excellence in Diabetes Care), Chennai, India; Blizard Institute, Queen Mary University of London, United Kingdom; Barts Health NHS Trust, London, UK

## Abstract

Type 2 diabetes (T2D) is a common and complex metabolic condition with significant heterogeneity within and across ancestries^1–4^. Compared with individuals of European ancestry (EUR), people of south Asian ancestry (SAS) have two to four-fold higher risk of T2D, develop the disease at younger ages and lower body mass index (BMI), and experience more rapid progression to complications^5–10^. Understanding the genetic basis of this is hindered by low representation of south Asians in genetic studies. Here, we perform an exome-wide association study of T2D in 13,674 cases and 41,024 controls from the Genes & Health study of British Pakistani and Bangladeshi individuals. We identify a novel rare variant in *HNF4A* – a canonical monogenic diabetes / MODY gene, in which missense variants would be expected to increase T2D risk. Surprisingly, *HNF4A* Pro437Ser is associated with a halved risk of T2D and reduced risk of diabetes-related complications but increased non-HDL cholesterol. We additionally characterise a T2D risk-increasing variant which is common only in South and East Asian ancestral groups (*GP2* Val429Met), which is associated with lower BMI and phenotypic and genetic markers of insulin deficiency. We validate our findings through replication in independent multi-ancestry cohorts, *in vitro* functional assays, and integration of proteogenomic analysis. These findings highlight how the study of under-represented populations can identify biological mechanisms associated with disease phenotypes enriched in those populations.

## MAIN

To explore the genetic basis of T2D in south Asians, we used REGENIE to test the association of common (minor allele frequency, MAF > 1%) and rare (MAF < 1%) variants with T2D in 13,674 cases and 41,024 controls from the Genes & Health study of British Bangladeshis and Pakistanis, after exclusion of 575 individuals based on predefined quality control criteria (Methods). Summary demographic information for included participants is given in **Table 1** and a participant flow diagram in **Fig S1**.

### An ancestry-enriched *HNF4A* variant underlies a protective association with Type 2 Diabetes

Rare variant aggregation analyses identified three significant gene-level associations with type 2 diabetes (T2D), (*MAP3K15*: predicted loss-of-function or predicted damaging missense (pLoF|pDM); *RNF19A*:pLoF; and *HNF4A*: pLoF|pDM (**Fig 1, S2-S4**, **Table S1**). Details on gene-testing and variants constituting each aggregate mask are provided in **Methods** and **Table S2**.

**Figure 1:**
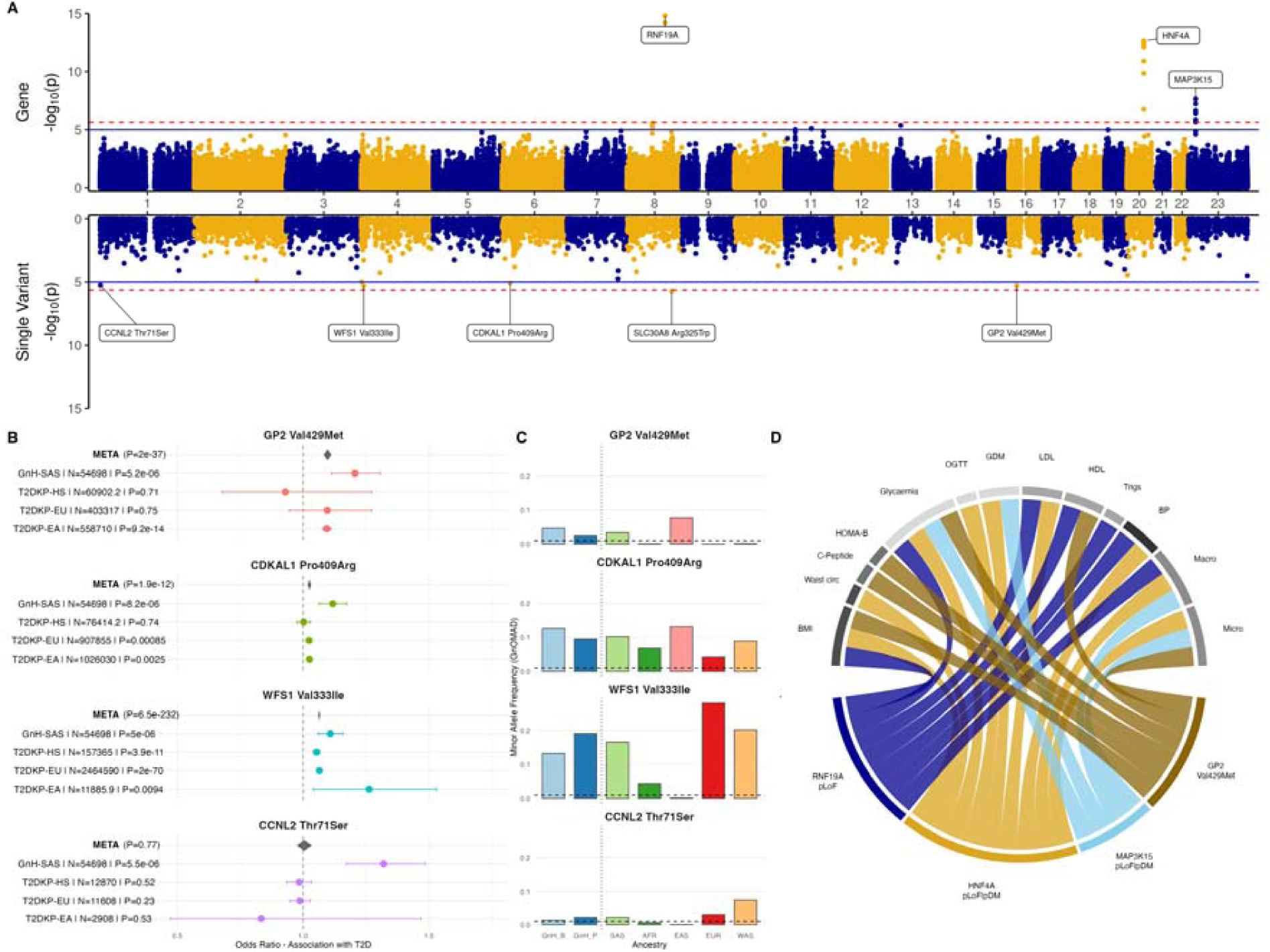
Summary of genetic discovery results for type 2 diabetes (T2D) association testing in the Genes & Health study, and associated phenotypes. **A:** Manhattan plot showing association with T2D for gene-based aggregates of rare variants with minor allele frequency (MAF) <0.01 (top panel) and single variant results for common variants with MAF > 0.01 (bottom panel). **B:** Trans-ancestry meta-analysis of single variant association with T2D. **C:** Bar plots showing corresponding MAF across gnomAD ancestries and in G&H sub-ancestries. **D:** Chord diagram showing association of selected results of interest from panel A with metabolically relevant phenotypes; a chord between a genetic variant / gene and phenotype indicates a significant association. GnH: Genes & Health. T2D-KP: type 2 diabetes knowledge portal. GnHB/P: Genes & Health Bangladeshis / Pakistanis; SAS: South Asian. HS: Hispanic. EUR: European. EAS: East Asian. AFR: African. WAS: West Asian (alternatively described as Middle Eastern). BMI: body mass index. Waist circ: waist circumference. Glycaemic: aggregate of HbA1c, fasting and non-fasting glucose. HOMA-B: homeostasis model assessment of beta cell function. OGTT: oral glucose tolerance test. GDM: gestational diabetes mellitus. LDL: low density lipoprotein cholesterol. HDL: high density lipoprotein cholesterol. Trigs: serum triglycerides. BP: systolic blood pressure. Macro: T2D macrovascular complications, composite of coronary artery disease, cerebrovascular disease, peripheral vascular disease. Micro: T2D microvascular complications, composite of nephropathy, neuropathy, and diabetic eye disease.

Consistent with previous reports ^11^, rare variants in *MAP3K15* were associated with reduced T2D risk, and a novel, nominal association was observed with increased risk of Transient Ischemic Attack in SAS-specific meta-analysis (**Fig S3A/B**). Trans-ancestry meta-analysis of the highly-conserved gene *RNF19A* showed consistent associations with increased T2D risk consistent increase in glycaemic markers, and nominally increased risk of ischemic heart disease (**Fig S3C/D, Table S3**). We identified further nominal associations of *RNF19A* with metabolic traits. Given prior evidence and limited interpretability, these exploratory findings were not pursued further.

In contrast, the *HNF4A* signal represented a novel direction of effect. Gene-based testing suggested a protective association (odds ratio (OR) = 0.56, 95% CI 0.48–0.67, P = 1.2 × 10⁻ ¹¹; **Fig 1A, S4A**), in contrast to the established role of deleterious *HNF4A* variants in monogenic diabetes.^12,13^ Leave-one-variant-out analyses demonstrated that this association was driven by a single missense variant, p.Pro437Ser (rs150776703), with no residual signal after its removal (**Fig 2A)**.

**Figure 2:**
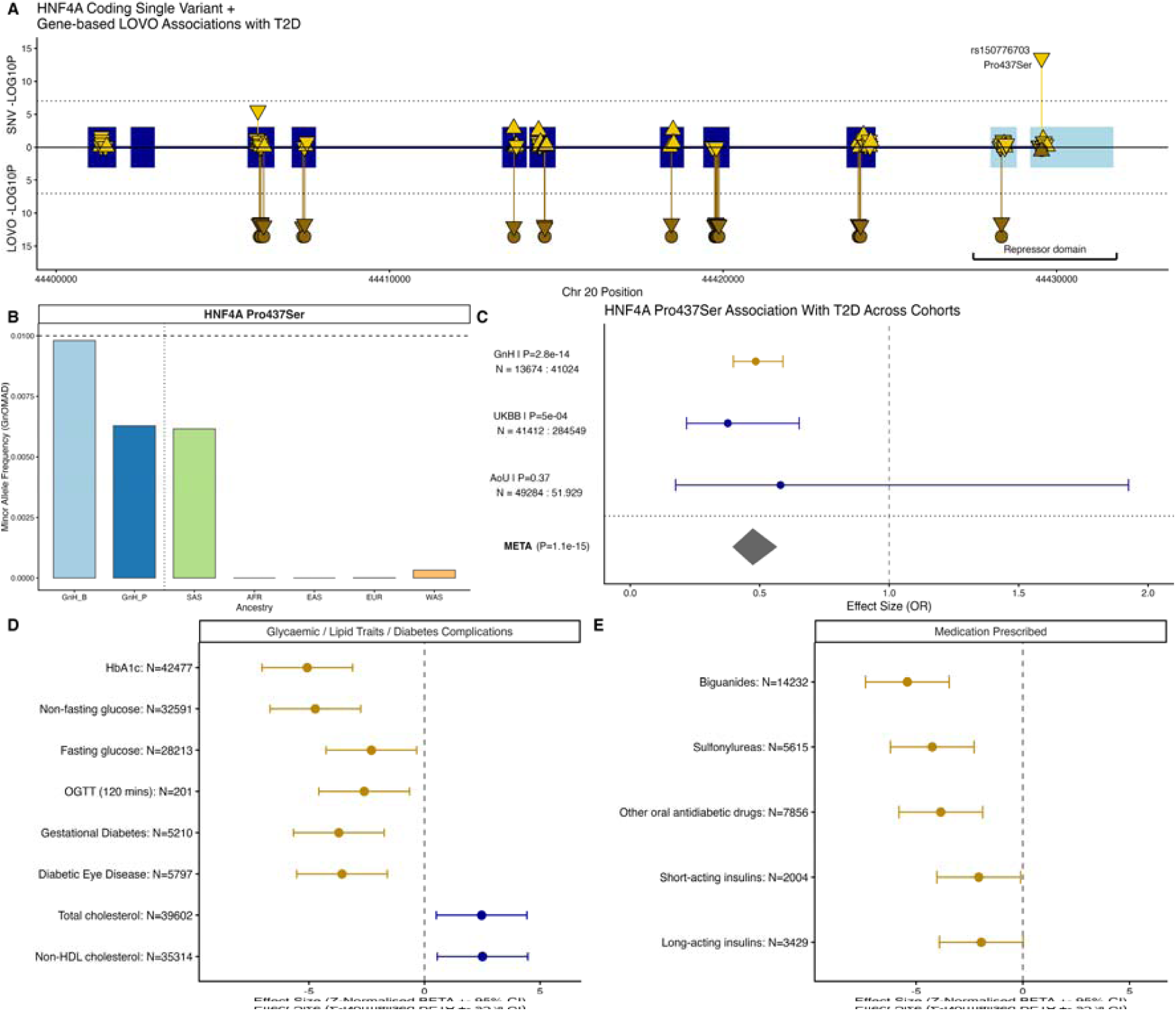
An ancestry-enriched HNF4A variant underlies a protective association with type 2 diabetes. **A:** Lollipop plot showing Chr20 genomic position relative to ENSEMBL canonical *HNF4A* exonic regions (dark blue rectangles; repressor exonic region light blue rectangles), overlaying single variant T2D association testing -LOG10P values for coding variants (top panel) and -LOG10P values from leave-one-variant-out (LOVO) analysis using the most significant mask in gene-burden testing (deleterious missense variants, MAF < 0.01) in the bottom panel. In this bottom panel, the LOG10P value for the mask without the named variant is plotted. Horizontal dotted lines represent exome-wide significance. The direction the triangle is pointing represents direction of effect, e.g. the labelled downward-pointing triangle Pro437Ser is risk-decreasing, **B:** Allele frequency of HNF4A Pro437Ser across ancestral groups; data are presented from gnomAD, or calculated allele frequencies in Genes & Health. **C:** Association of HNF4A Pro437Ser with type 2 diabetes in Genes & Health, with replication in UK Biobank (UKBB) and All of Us (AoU), and meta-analysis across cohorts. **D/E:** Z-normalised betas and 95% confidence intervals from multivariable regression models adjusted for age, age^2^, sex, and genetically-inferred ancestry, describing the association of *HNF4A* Pro437Ser with (D) clinical outcomes in the electronic health record and (E) use of glucose-lowering therapies. For binary traits (e.g. panel E) the BETA represents the log(odds ratio). HbA1c, glycated haemoglobin; OGTT, oral glucose tolerance test. GnHB/P: Genes & Health Bangladeshis / Pakistanis; SAS: South Asians: AFR: African; EAS: East Asian; EUR: European; WAS: West Asian (Middle Eastern).

Single-variant analysis confirmed a strong protective association between p.Pro437Ser and T2D (OR = 0.48, 95% CI 0.40–0.59, P = 2.8 × 10⁻¹ □). The variant is enriched in South Asian populations (gnomAD minor allele frequency = 0.008) and rare or absent in other ancestries **(Fig 2B)**. Replication in UK Biobank and All of Us demonstrated consistent direction of effect, with meta-analysis providing strong support for association (OR = 0.47, P = 1.5 × 10⁻¹□) (**Fig 2C**). Further details on fine-mapping are provided in the **supplementary results and Table S4.**

### *HNF4A* Pro437Ser is associated with favourable glycaemic traits but higher lipid levels

Carriers of *HNF4A* Pro437Ser exhibited improved glycaemic profiles, lower HbA1c (β= −2.23 mmol/mol, 95% CI −3.10 to −1.37; P = 4.0 × 10⁻□) and lower non-fasting glucose levels (β = −0.44 mmol/L, 95% CI −0.63 to −0.26; P = 2.4 × 10⁻□), alongside reduced risk of diabetic eye disease (OR = 0.96, 95% CI 0.94–0.98; P = 3.0 × 10⁻ □) and gestational diabetes (OR = 0.95, 95% CI 0.92–0.98; P = 0.005 (nominal significance)) (**Fig 2D; Table S5).** Congruently, variant carriers had lower odds of lifetime use of any glucose-lowering therapies such as biguanides (OR = 0.92, 95% CI = 0.91-0.94, p = 1.1 x 10^-8^(**Fig 2E**; **Table S4**). In contrast, the variant was associated with higher total cholesterol (beta = 0.21, 95% CI 0.12–0.32, P = 1.3 × 10⁻□) and non-HDL cholesterol (beta = 0.09, 95% CI 0.02–0.17, P = 0.009 (nominal significance)), indicating divergent metabolic effects (**Fig 2D, Table S5**).

#### The protective effect of *HNF4A* is variant-specific rather than domain-driven

Given the location of Pro437Ser within the *HNF4A* repressor domain^14^ **(Fig 2A)**, and most HNF4A-MODY causing variants lie upstream of this^15^ we assessed whether variant effects differed by protein domain. While variants in non-repressor regions were associated with increased T2D risk in Genes & Health (OR = 1.41, 95% CI = 1.05 – 1.88, p = 0.02), consistent with monogenic diabetes, the apparent protective signal within the repressor domain was entirely attributable to Pro437Ser. Exclusion of this variant abolished the association in Genes & Health (OR = 0.85, 95% CI = 0.53-1.36, p = 0.50). Meta-analysis of repressor domain associations with T2D in the absence of Pro437Ser in UKBiobank (n= 459,903), and AMP-T2D (Accelerating Medicines Partnership in Type 2 Diabetes, n = 20,074). demonstrated increased risk of T2D (OR = 1.51, 95% CI = 1.12 – 2.05, p = 0.003). These findings suggest that the protective effect of HNF4A Pro437Ser is likely variant-specific rather than a general property of repressor domain variation (**Fig 3A; Table S6-8; Supplementary Results**).

**Figure 3.**
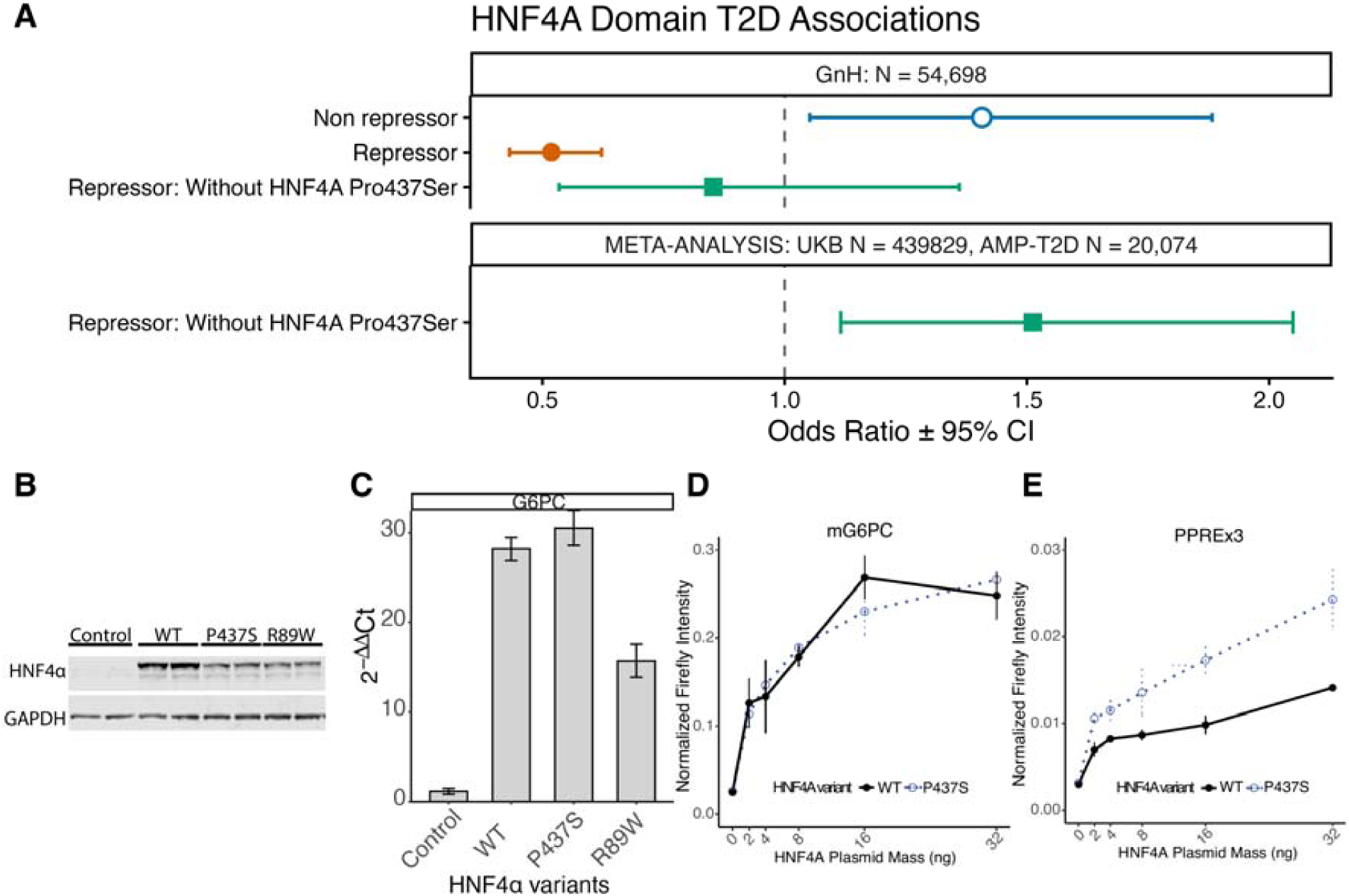
Variant-specific effects of HNF4A Pro437Ser and target-dependent transcriptional activity. **A.** Gene-burden analyses of rare missense variants (MAF < 0.01) in HNF4A, stratified by protein domain. Variants in the non-repressor domain were associated with increased T2D risk, consistent with monogenic diabetes, whereas the apparent protective association in the repressor domain was driven by inclusion of Pro437Ser and was not observed after its exclusion. Findings were consistent across Genes & Health and external datasets (UK Biobank and AMP-T2D; see Supplementary Information). **B.** Western blot of Huh7 cells overexpressing wild-type (WT), Pro437Ser (P437S), or Arg89Trp (R89W) HNF4A, with GAPDH as loading control. **C.** Expression of the endogenous HNF4A target gene G6PC in Huh7 cells, showing comparable activity of WT and Pro437Ser, and reduced activity of Arg89Trp. **D.** Dose-dependent transactivation of a mouse G6PC promoter-driven luciferase reporter by HNF4A variants in HEK293 cells. **E.** Dose-dependent transactivation of a synthetic PPRE×3 minimal promoter luciferase reporter by HNF4A variants in HEK293 cells, showing modestly increased activity of Pro437Ser relative to WT. Reporter assays were performed across independent biological replicates, with firefly luciferase activity normalised to Renilla.

#### HNF4A Pro437Ser exhibits target-dependent differences in transcriptional activity

To assess functional consequences, we evaluated the transcriptional activity of *HNF4A* Pro437Ser in cellular assays. Wild-type (WT), Pro437Ser, and a known loss-of-function MODY variant (Arg89Trp)^16^ were overexpressed in Huh7 hepatocyte-derived cells, with robust protein expression confirmed by western blot (**Fig 3B**). WT HNF4A induced expression of the endogenous target gene G6PC (28-fold increase), while Pro437Ser showed comparable activity, and Arg89Trp demonstrated reduced transactivation, consistent with impaired function **(Fig 3C)**.

We next performed dual-luciferase reporter assays to compare transcriptional activity across promoter contexts. HEK293 cells were co-transfected with increasing amounts of *HNF4A* expression constructs alongside either a mouse G6PC promoter reporter **(Fig 3D)** or a synthetic PPRE×3 minimal promoter containing three consensus DR1 motifs **(Fig 3E)**. Both reporters exhibited dose-dependent activation across independent biological replicates; however, their response profiles differed markedly. The mG6PC promoter, which contains non-consensus *HNF4A* motifs and requires additional co-factors for full transcriptional activation^17^, showed a nonlinear activation curve with no clear separation between WT and Pro437Ser across the tested range. In contrast, the PPRE×3 reporter displayed a near-linear response, with modestly increased transactivation by Pro437Ser relative to WT.

Together, these findings indicate target-dependent differences in transcriptional activity, whereby the effect of Pro437Ser is more evident in simplified promoter contexts with optimal binding motifs and minimal cofactor constraints, but attenuated at native metabolic promoters, where transcription is constrained by low-affinity motifs and the absence of required cofactors.

### *GP2* variation highlights a β-cell–related pathway of type 2 diabetes risk

Single-variant analyses identified several coding variants associated with type 2 diabetes, including signals at previously characterised loci (for example, *SLC30A8*, *CDKAL1* and *WFS1*), which were not pursued further. We therefore focused on a missense variant in *GP2* (p.Val429Met), which is enriched in East (MAF 0.07) and south Asian (0.03) populations and has not been extensively characterised **(Fig 1B/C)**. Within south Asian sub-populations the variant is more common amongst Bangladeshis (0.047) compared to Pakistanis (0.025) and is ultra-rare in other populations. This is consistent with substantial allele frequency differentiation across populations (**Supplementary Results**).

In trans-ancestry meta-analyses, *GP2* Val429Met was consistently associated with increased risk of T2D (rs78193826 C>T; Val429Met; OR = 1.09, 95% CI = 1.08-1.10, p = 2.04 x 10^-37^ **(Fig 1B).** This signal has previously been reported in East Asian populations (AGEN consortium and Biobank Japan) but has not been extensively characterised, nor reported in other ancestries^18,19^.

The effect of Val429Met on glycaemic traits was evaluated in trans-ancestry meta-analysis, using data from G&H, MDRF and EAS cohorts in T2DKP^20^ and insulin response from the MAGIC consortium^21^. Val429Met was significantly associated with higher fasting glucose (ß= 0.03 mmol/L, 95% CI 0.02 – 0.04, p = 7.4 x10^-11^) and nominally with lower 2hr C-peptide levels (ß = -0.08 nmol/L, 95% CI = -0.13 - -0.03, p = 0.002, n=14,704) and lower BMI (-0.009 kg/m^2^, 95% CI = -0.002 - -0.016, p = 0.02) (**Fig 4A, Fig S5, Table S9).** In contrast, the variant was not associated with measures of insulin resistance including HOMA-IR and Triglyceride:HDL ratio, suggesting a role in insulin secretion pathways.

**Figure 4.**
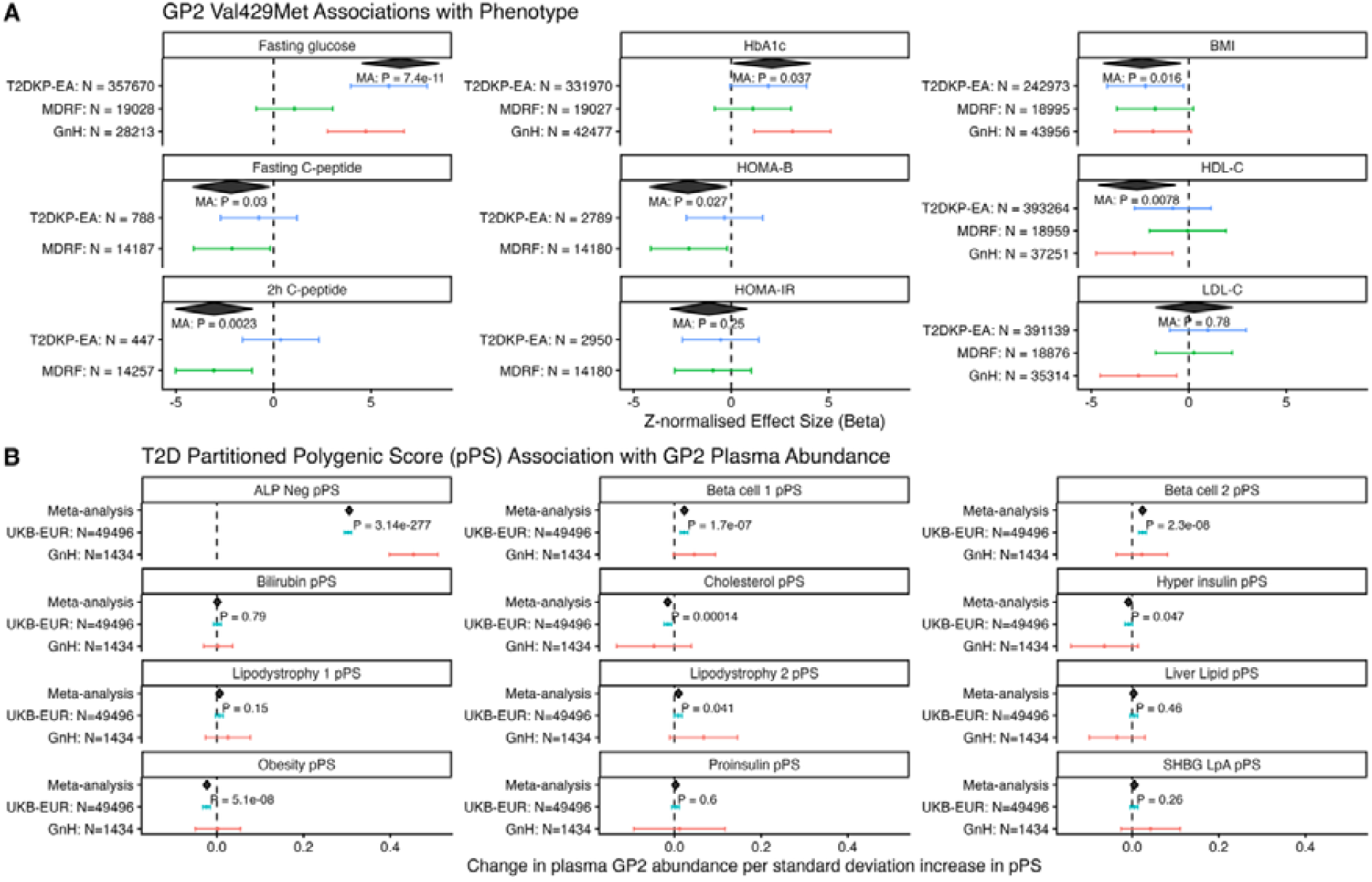
Characterisation of GP2. **(A)** Trans-ancestry meta-analysis of GP2 Val429Met variant association with glycaemic, beta-cell, and lipid traits across South and East Asians in Genes & Health, the Madras Diabetes Research Foundation (India), and East Asian summary statistics from the Type 2 Diabetes Knowledge Portal. Traits are Z-transformed to allow presentation on the same scale and are thus plotted without units. Not all traits were available in all datasets. **(B)** Association of GP2 circulating protein abundance in plasma, measured using the Olink platform in Genes & Health (GnH), n = 1434, and UK Biobank Europeans (UKB_EUR), n = 49,496, with pathway-specific partitioned polygenic scores (pPS) for type 2 diabetes taken from Smith et al. Results are plotted as the beta per standard deviation increase in each pPS, estimated from a multivariable regression model adjusted for age, sex and ancestry. For example, increases in genetic risk of type 2 diabetes mediated via insulin sensing (Beta Cell 1) or insulin deficiency (Beta Cell 2) endotypes are associated with increased GP2 plasma abundance. ALP Neg: ALP negative; SHBG: sex hormone binding globulin.

Consistent with this, GP2 is predominantly expressed in the pancreas and more specifically pancreatic acinar cells, where it forms part of the zymogen granule membrane, with minimal expression in islets (**Fig S6A/B)**^22–24^. Proteomic analyses showed increased circulating GP2 levels associated with *GP2* Val429Met on one platform (Olink), **(Fig S6C)**, and integration with a human plasma proteome atlas^25^ supported the pancreas as the primary tissue of origin for circulating GP2 (**Fig S6D).** However, changes in GP2 abundance were not observed consistently across assays (non-significant association on SomaScan platform, **Fig S6C),** raising the possibility that missense variation altering protein structure could interfere with one (or both) assays through binding or epitope artefacts with disruption of antibody or aptamer binding.

To further contextualize the role of GP2 within the heterogeneous genetic architecture of T2D, we examined associations between previously defined T2D genetic endotypes called partitioned polygenic scores (pPS)^26,27^ and plasma *GP2* levels in individuals of European ancestry from UK Biobank and in G&H (Olink), irrespective of diabetes status. Curation of Olink data have been described previously^28^.

Trans-ancestry proteogenomic meta-analysis demonstrated that higher genetic burden of T2D related to β-cell dysfunction was associated with increased plasma GP2 levels (beta-cell 1 (insulin sensing): beta = 0.02, 95% CI = 0.012 – 0.028, p = 1.74 × 10⁻ □; beta-cell 2 (insulin secretion): beta = 0.02, 95% CI = 0.013-0.028, p = 2.35 × 10⁻ □)(**Fig. 4B).** Conversely, greater obesity- and lipid-related genetic risk was associated with lower GP2 levels. Increased plasma abundance of GP2 in the context of greater genetic risk of beta-cell dysfunction could, for example, indicate increased cell destruction.

Together, these results extend previous association signals at the *GP2* locus by integrating genetic, phenotypic and proteomic data, supporting a role in beta-cell–related pathways of type 2 diabetes risk. Caution is required in interpreting proteomic results in the absence of independent verification of GP2 abundance-related findings on an alternate platform to Olink, given discordance with SomaScan in Genes & Health, where both platforms are available.

## Discussion

In this exome-wide association study of type 2 diabetes in a south Asian population, we identify an ancestry-enriched coding variant in *HNF4A* associated with reduced disease risk and extend the characterisation of variation at the *GP2* locus, illustrating complementary mechanisms contributing to diabetes susceptibility. These findings demonstrate how studying populations with distinct genetic architectures can reveal biological insights that are difficult to access in other datasets but are relevant to general disease biology.

The protective effect of *HNF4A* Pro437Ser contrasts with the established role of deleterious variants in this gene^13,29^ in monogenic diabetes and is not shared across other variants, indicating a variant-specific effect. Functional data support target-dependent differences in transcriptional activity, highlighting the complexity of transcription factor perturbation. Notably, the variant is associated with improved glycaemic traits but adverse lipid profiles, suggesting metabolic trade-offs similar to those reported for related transcription factors such as HNF1A^30^, and indicating that further mechanistic characterisation will be required.

At the *GP2* locus, integrative genetic and proteomic analyses support a role in β-cell–related pathways, in the absence of obesity-related risk of type 2 diabetes. This resembles the phenotype of young, and lean-onset diabetes observed frequently in South Asian populations^27,31,32^.

Together, these findings illustrate how ancestry-enriched genetic variation can resolve distinct components of diabetes biology and inform future mechanistic and therapeutic studies.

## Methods

### Study design

We performed a type 2 diabetes (T2D) case-control exome-wide association study using participant data from the Genes & Health (G&H) study^33^ based in the UK, focusing separately on rare variants with minor allele frequency (MAF) < 1% and common variants with MAF > 1%. For significant results passing Bonferroni correction (p < 9.35 x 10^-8^), we investigated associations with metabolically relevant phenotypes in G&H. We replicated and meta-analysed T2D associations using data from UK Biobank^34,35^, All of Us^36^, and the Type 2 Diabetes Knowledge Portal (T2DKP)^20^, a live, open access resource collating and meta-analysing summary statistics across ancestries for type 2 diabetes and related traits. We included 54,698 individuals (13,674 type 2 diabetes cases and 41,024 controls, 575 excluded) in the exome-wide association study in Genes & Health; further details are given in **Figure S1.** An overview of study design and features in line with the STREGA reporting guideline^37^ is presented in the accompanying STREGA checklist.

### Setting

Genes & Health is a long-term community-based population cohort, funded to recruit 100,000 British Bangladeshi and British Pakistani individuals in the UK aged 16 and over, linking genetic testing to routine, longitudinal, electronic health record (EHR) data^38^. To date, Genes & Health has recruited ∼75k volunteers, among whom 55,273 have whole exome sequencing data, and 51,170 have imputed genotype data, with linkage to primary and secondary care clinical diagnostic codes, laboratory test results, and prescribing data.

In addition to using publicly available summary statistics, we replicated and meta-analysed findings in a separate setting, the MDRF biobank in India^39^. This is a national-level, multicentre, clinic-based disease registry, combining phenotypic data with genetic testing in a nationally-representative sample of individuals with T2D (n = 19,898) and healthy controls (n = 4,711).

### Participants

#### Inclusion criteria

We used no specific inclusion criteria beyond those of the Genes & Health study.

#### Exclusion criteria

We excluded from analyses individuals diagnosed with type 2 diabetes before 18 years of age, and those with a clinical code of type 1 diabetes, maturity onset diabetes of the young (MODY), neonatal diabetes, secondary diabetes (e.g. drug-induced diabetes), or clinical conditions strongly predisposing to secondary diabetes (e.g. cystic fibrosis, pancreatectomy). Clinical code lists used for exclusion criteria are provided in **Table S10**.

#### Case definition (Genes & Health)

We defined type 2 diabetes as an entry of type 2 diabetes in the National Diabetes Audit (NDA)^40^, an annual England-wide registry of type 2 diabetes patients. Because NDA data are not yet available from 2023 onwards in the G&H data linkage, we also defined as type 2 diabetes individuals with a first clinical code of type 2 diabetes in the EHR from 2023 onwards. For cases, we defined “age at diagnosis” as the age at the earliest clinical code of type 2 diabetes in the EHR.

#### Control definition (Genes & Health)

We defined controls as individuals with no record of type 2 diabetes in the NDA, and with no records of type 2 diabetes in the EHR at any time.

#### Case & Control Definitions (MDRF)

The MDRF clinical cohort comprises of data extracted from electronic medical records (EMR) of Dr Mohan’s Diabetes Specialities Centre, a large tertiary diabetes care network with over 50 centres across 32 cities in 8 states of India. For individuals who underwent GWAS, demographic variables including sex, age, diabetes type (e.g. type 2 diabetes, MODY, etc.), age at diagnosis and duration of diabetes as well as anthropometric details, biochemical data were extracted. Fasting and stimulated (post-breakfast) serum C-peptide concentrations were quantified using an electrochemiluminescence immunoassay on the Elecsys 2010 analyser (Hitachi). Venous blood samples were obtained after an overnight fast of >8 hours, followed by collection of a postprandial sample 90 minutes after consumption of a standardized mixed breakfast providing approximately 250 kcal. HOMA-B is derived using C-peptide serves as an index of pancreatic beta cell secretory function and is calculated from fasting plasma C-peptide and plasma glucose concentrations using the Homeostasis Model Assessment (HOMA) calculator (University of Oxford, UK), as previously described. T2D was defined by the absence of ketosis, good beta-cell functional reserve as evidenced by stimulated C-peptide (>0.6 pmol/ml), absence of pancreatic calculi (on X-ray abdomen) and good response to oral hypoglycaemic agents for more than two years.

### Variables

#### Exposures

We performed genetic association testing using both whole exome sequencing (exome-wide association study, ExWAS) and imputed genotype data (genome-wide association study, GWAS).

#### Outcomes

We defined one primary outcome: type 2 diabetes state. This was a binary variable used in regression-based association testing.

We defined a larger number of secondary outcomes (27 quantitative, 8 binary disease and 5 binary medication prescription traits, **Table S2).** We examined the association of these traits with genetic exposures of interest passing the Bonferroni-corrected threshold. Further detail is provided in the “statistical methods” section below and the **supplementary methods**.

#### Confounders

We adjusted analyses for confounders typically included in multivariable genetic association models: age at recruitment, age at recruitment squared, sex, and the first 20 genetic principal components.

##### Sex-stratified analysis

For each analysis, we additionally ran sensitivity sex-stratified models (defined as the presence or absence of a Y chromosome) to explore the possibility of sex-specific effects (**Table S4)**.

### Data sources and measurements

#### Sample Whole Exome Sequencing

Exome sequencing was performed on 55,273 individuals by the Broad Institute using Twist Reagents and Illumina 150bp PE Novaseq-6000 sequencing, mapped to hg38 to produce crams as previously described^41^. Crams with less than 85% of bases at >20x coverage were excluded. UMAP was performed on the first 7 genetically determined principal components to define Pakistani and Bangladeshi ancestry individuals, which correlated highly with 1000 genomes population references; 198 outlier samples not corresponding to Pakistani or Bangladeshi ancestry were not included in analyses.

*Variant QC* was performed using a random forest model trained on a set of chromosome 20 genotype-corresponding variants identified as true positives satisfying the following criteria: sites discovered in 1000 Genomes with high confidence; SNVs present in the Omni 2.5 genotyping array and found in 1000 Genomes; indels present in Mills & Devine data^42^; HapMap3 SNVs and Indels. False positive variants were defined as failure at any of: normalized quality by depth (QD) score < 2; Fisher strand bias > 60; mapping quality (MQ) < 30. The random forest model included 12 features; the highest relative importance were QD (weighted importance = 0.41), mean heterozygous allele balance (0.34), strand odds ratio(0.09), MQ (0.05), MQ_RankSum (0.04) multiallelic site flag (0.03), and number of alternate alleles at a site(0.05). Further details can be found in the supplemental information of ^43^.

#### Sample Genotyping (for fine-mapping)

We used TOPMed-r3 imputed genotype information from Illumina Infinium Global Screening Array v3 (with additional multi-disease variants), performed as previously described in 51k individuals^27^. We excluded variants with low INFO scores below 0.7. We performed a matched genome-wide association study using the same T2D phenotype as defined above, in 45,634 individuals with both WES and imputed genotype data. We used these summary statistics for fine-mapping variants of interest identified in our ExWAS.

### Statistical methods

#### Exome-wide association study (ExWAS) – gene-based testing

We used REGENIE v4.0 to examine the association of 534,315 gene-based “masks” previously defined^41^ as sets of variants below three MAF thresholds (0.01, 0.001, 0.0001) or singleton, with specific functional annotations (LOFTEE_High_confidence_predicted loss_of_function (pLoF), predicted damaging missense variants (CADD > 20 & PolyPhen > 0.466 and SIFT<0.05, pDM), other missense variants (causing amino acid substitution but not meeting the above criteria), and synonymous variants); full definitions are given in **Table S1**. We defined gene testing exome-wide significance as a Bonferroni-corrected p value of p < 9.35 x 10^-8^. (ie, 0.05 / 534,315 tests). Association models were adjusted for age at recruitment, age at recruitment squared, sex, and the first 20 genetic principal components.

Models were run as REGENIE v4.0 “binary trait” models with approximate Firth correction below p = 0.05. For Step 1, we used an LD-pruned subset of common (MAF > 0.01) genotyped markers created using the plink command “—indep-pairwise 1000 100 0.9 (n_variants = 78,354). For Step 2, we employed the “leave one chromosome out (LOCO)” method to avoid proximal contamination. All 54,698 included participants were included in step 1 and step 2.

We performed rare variant association testing across 18,956 genes by aggregating rare variants with MAF < 1% into functional categories (i.e. masks, as previously described^43^ and in **Table S1)** and testing the association of these aggregates with T2D. After correcting for multiple-testing (p<9.35 x 10-8, methods)

For *HNF4A*, identified as a key gene of interest, we additionally performed leave-one-variant-out analysis using the “LOVO” option in REGENIE. In addition, we constructed bespoke masks defined using the variant categorisation above, but stratified by whether each variant lay within an *HNF4A* repressor or non-repressor domain, to explore whether the effects of identified variants differed by proposed exonic repressor-based location. Replication of this approach in UK-Biobank and AMP-T2D (Accelerating Medicines Partnership in Type 2 Diabetes) are described in the **supplementary methods.**

#### Domain-specific burden testing

Having identified a risk-reducing variant in the repressor domain of *HNF4A*, we sought to explore whether other missense variants in the gene’s repressor domain were associated with reduced risk of T2D; ie, to explore whether the negative association reflected a repressor domain effect, or a variant-specific effect. In addition to GnH analyses, we included 8,875 T2D cases and 11,199 controls from the AMP-T2D GENES (55k) cohort^44^. 36,637 T2D cases and 403,192 controls from the UK Biobank, following the T2D definition created previously^44^. Variants were annotated using VEP v110.1 with LOFTEE v1.0.4 plug-in and dbNSFP v4.4a plug-in. We only selected variants in the transcript ENST00000316099 within the “repressor” F domain (chr20:44,424,233-44,424,254; chr20:44,428,336-44,428,458; chr20:44,429,522-44,429,661). We removed rs150776703 (20:44429549:C:T) from the analysis. We grouped the selected variants into 16 masks proposed by a previous study^45^, and two loss-of-function masks (high-confidence predicted by LOFTEE, MAF < 1% and MAF < 0.1%). We then conducted burden tests in the UK Biobank and AMP-T2D GENES cohorts separately using REGENIE. For step 1, we used the following parameters: “--bsize 200 –bt”. For step 2, we used the following parameters: “--minMAC 1 --bsize 200 --aaf-bins 0.5,0.01,0.001 --build-mask sum --bt --af-cc --firth --firth-se --write-mask-snplist”.

In both cohorts, we only kept the burden results of masks where cumulative alternate allele counts were greater than 20. We next meta-analyzed the results from two cohorts for each remaining mask using the inverse-variance schema in METAL. Finally, we applied the “minimum p-value test”^1^ to aggregate the p-values across all masks.

#### Exome-wide association study (ExWAS)– single variant

We examined the association of 25,326 genetic variants with MAF > 1% in 54,698 participants using REGENIE v4.0. Because of high inbreeding coefficients among variants with MAF ∼ 0.50, we cautiously excluded 5048 variants in the window 0.46<MAF<0.54, and restricted analysis to single nucleotide polymorphisms. The methods for this analysis were otherwise identical to the GWAS methods above. We defined exome-wide significance as a Bonferroni-corrected p value of p < 1.97x 10^-6^ (ie, 0.05 / 25,326 tests).

#### Association testing with metabolic traits and diabetes-related complications

We explored the association of variants of interest with 40 binary and quantitative traits (**Table S11)** in the Genes & Health study defined using robust, open-source pipelines using multivariable regression models adjusted for age at study recruitment, sex, ancestry, and the first 20 principal components. These include binary trait phenotypes defined as ICD10 codes, including codes mapped from SNOMED (used in Genes & Health primary care data) to ICD10 (which are used for Genes & Health secondary care data), as described previously^27^. For T2D microvascular complications (diabetic eye disease, neuropathy, and nephropathy) and macrovascular complications (cardiovascular disease, cerebrovascular disease, peripheral vascular disease) we defined only incident complications after the date of T2D diagnosis as T2D complications.

For quantitative traits, we explored the association of variants of interest with lifetime median values in all included participants, agnostic to T2D status, as described previously^46^.

We additionally explored the association of variants of interest with traits related to insulin resistance at the time of diagnosis in 13,468 type 2 diabetes cases. We defined traits at the time of diagnosis as a reading within 1 year (either before or after) the time of earliest date of type 2 diabetes diagnosis in the EHR, as described previously. Where multiple measures were available within this window, we selected the value closest in time to diagnosis data. Associations were reported from regression models adjusted for age at study recruitment, sex, ancestry, and the first 20 principal components.

We explored the association of genetic variables with prescription of diabetes-controlling medication in a subset of 43,076 individuals for whom primary care prescribing data was available, defined by British National Formulary (BNF) chapter.^47^ We defined exposure to a diabetes-controlling medication as ever having been prescribed a medication from a BNF chapter, and explored association using adjusted multivariable regression models as described above for other traits. We included 6 medication-related outcomes: biguanides, sulfonylureas, “other antidiabetic medication” (including SGTL2i and GLP1-agonists), short-acting insulins, and long-acting insulins.

We present results from this targeted phenome association testing as significant if P < 0.05 / 40 (= P < 0.00125); we describe results with 0.05 > P > 0.00125 as nominally significant.

After establishing that the gene-based signal for one discovery result (*HNF4A)* was driven by a single variant, we additionally examined phenotypic associations of that single variant, using the same approach as described above.

#### Meta-analysis

We meta-analysed T2D association results for type 2 diabetes case-control status and phenotypes identified at nominal significance (p<0.05) in Genes & Health trait association testing with replication results from UKBiobank and AllofUs; the association testing producing these statistics is described in^35,44^.

For metabolic trait associations from gene-based tests, we meta-analysed with burden summary statistics from UK Biobank available through the AstraZeneca PheWAS portal^34^. This resource provides gene-burden summary statistics across multiple masks, stratified by ancestry; we selected the most closely aligned mask to the Genes & Health discovery mask for each gene (e.g., protein-truncating variants in UKB for high-confidence predicted loss of function variants in GnH). Because not all REGENIE gene masks produce a direction of effect with associated beta and standard error (e.g. GENE-P), for phenotypes with nominal significance (p < 0.05) in internal association testing but no associated beta in the most significant mask*test combination, we selected the most significant gene-burden mask*test combination within Genes & Health with a direction of effect, measured as the smallest p value.

We meta-analysed single variant associations for phenotypes of interest identified at nominal significance (p<0.05) in Genes & Health trait association testing with publicly available summary statistics aggregated via the T2DKP. Where possible, we meta-analysed with traits from MDRF, including meta-analysis between MDRF and the T2DKP for traits not included in the “discovery” sample Genes & Health, and particularly for traits related to HOMA estimates of insulin resistance and production. Meta-analysis was performed using fixed-effect models with the R package metafor v4.6-0.

### Functional characterization of HNF4A variant

#### qRT-PCR analysis of transcriptional activities across HNF4A variants

pTwist-CMV-puro expression vectors were generated by Twist Bioscience (San Francisco, CA, USA) for the following *HNF4A* variants: wild-type (WT, Uniprot isoform P41235-1, NM_000457.6), Pro437Ser (NM_000457.6:c.1309C>T), and Arg89Trp (NM_000457.6:c.265C>T), a confirmed loss of function variant^16^. Each construct included a C-terminal Myc tag. The *HNF4A* constructs were transfected into WT HUH7 hepatocytes using TransIT-LT1 transfection reagent (#MIR2304, Mirus Bio, Madison, WI, USA) following the manufacturer’s protocol. Variant overexpression was confirmed 24 hours after transfection using Western blotting with primary antibodies against c-Myc (#3-2500, Invitrogen, Thermo Fisher Scientific, Waltham, MA, USA) or HNF4A (#3113SCell Signaling Technology, Danvers, MA, USA) in three independent replicates. Expression of the HNF4A transcriptional target *G6PC* was assayed in HUH7 cells transiently transfected with an empty control vector (pLenti SpBsmBI sgRNA Puro (#62207, Addgene, Watertown, MA, USA)) or HNF4A WT, Pro437Ser, or Arg89Trp Myc-tagged constructs after 24 hours by qPCR as we have published previously^48^. The following primers were used for *G6PC:* FWD 5’- GCTGTGATTGGAGACTGGCTCA-3’, REV REV 5’-CAGGTTGGGGGTCAGTACC-3’; and the housekeeping control gene *GAPDH*: FWD 5’- CATCTTCTTTTGCGTCGCCA-3’, REV 5’- TTAAAAGCAGCCCTGGTGACC-3’.

#### Dual-luciferase analysis of transactivation activities across HNF4A variants

HEK293 cells were co-transfected with either a mouse G6PC promoter–driven firefly luciferase reporter (10 ng/well)^16^ or a PPRE×3-TK-Luc reporter (10 ng/well), together with a Renilla luciferase plasmid (10 ng/well) (pRL-SV40 for mG6PC or pGL4.75 for PPRE×3) as a transfection efficiency control. A 2-fold serial dilution of *HNF4A* variant expression plasmids (0–32 ng per well in a 96-well format) was used to assess dose-dependent transactivation. To maintain a constant total DNA amount across all conditions, a neutral pLenti-SpBsmBI-Puro backbone plasmid was added as filler DNA, and the final DNA mass and TransIT-LT1 reagent volume were kept constant for all conditions. Cells were harvested 48 h post-transfection, and firefly and Renilla luciferase activities were measured using the Dual-Luciferase Reporter Assay System (#E1910, Promega, Madison, WI, US) on a Tecan Spark multimode plate reader (Tecan Group Ltd., Männedorf, Switzerland). Firefly luciferase readings were normalized to Renilla to control for transfection efficiency. All experimental conditions were performed in three biological replicates.

#### Population genetic analysis

To test how likely it is to observe differenced in allele frequency among populations in target variants, we simulated the neutral evolution of 10,000 independent SNPs under a previously proposed joint demographic model for Europeans, South Asians and East Asians with 1,000 sampled individuals each using the *ms* software. We then retained simulations that yielded European allele frequencies lower than 0.2% and calculated the empirical rank p-value as the proportions of simulations generating frequencies in South and East Asians greater than the ones observed.

## Data Availability

Some data contained in this manuscript are publicly available at https://t2d.hugeamp.org/ and https://www.azphewas.com/.
Genes & Health data is available to bona fide researchers submitting an application to https://www.genesandhealth.org/researchers/apply-for-access/.
Information on applying to access MDRF data is available at https://www.mdrf.in/department/Data%20Management.html

## Acknowledgements

SH is funded by Wellcome Health Advances in Underrepresented Populations (HARP) Doctoral Fellowship 227532/Z/23/Z. VB is funded by the Wellcome Trust PhD programme - health data in practice: human-centred science (218584/Z/19/Z). ARM, AB and S Hu are supported by I01BX006293 from Veterans Affairs Office of Research and Development. DS and SF are funded by the Tackling Multimorbidity at Scale Strategic Priorities Fund programme (MR/W014416/1) delivered by the Medical Research Council and the National Institute for Health Research in partnership with the Economic and Social Research Council and in collaboration with the Engineering and Physical Sciences Research Council. IB and SF are funded by a Wellcome Discovery Award (227897/Z/23/Z) and IB acknowledges support from the National Institute for Health and Care Research Exeter Biomedical Research Centre. The views expressed are those of the authors and not necessarily those of the NIHR or the Department of Health and Social Care. GK & HCM’s involvement in this research was funded in part by Wellcome Trust (grant no. 220540/Z/20/A, “Wellcome Sanger Institute Quinquennial Review 2021–2026”). MKS is supported by Barts Charity grants MGU0504 and G-002995. Analyses in UKBiobank by AYD were performed under study accession number 53639. We acknowledgement the support of Indian Council of Medical Research (ICMR) for setting up the MDRF – Biobank.

J.M.M. is supported by American Diabetes Association grant #11-22-ICTSPM-16 and by NHGRI U01HG011723, by the National Institute Of Diabetes And Digestive And Kidney Diseases of the National Institutes of Health under Award Number R01DK137993, R01DK140545 and U01 DK140757, AMP CMD award from RFP 6 from the Foundation for the National Institutes of Health, and a Medical University of Bialystok (MUB) grant from the Ministry of Science and Higher Education (Poland). This work is supported by the Novo Nordisk Foundation (NNF21SA0072102).

Genes & Health is/has recently been core-funded by Wellcome (WT102627, WT210561), the Medical Research Council (UK) (M009017, MR/X009777/1, MR/X009920/1), Higher Education Funding Council for England Catalyst, Barts Charity (845/1796), Health Data Research UK (for London substantive site), and research delivery support from the NHS National Institute for Health Research Clinical Research Network (North Thames). We acknowledge the support of the National Institute for Health and Care Research Barts Biomedical Research Centre (NIHR203330); a delivery partnership of Barts Health NHS Trust, Queen Mary University of London, St George’s University Hospitals NHS Foundation Trust and St George’s University of London

Genes & Health is/has recently been funded by Alnylam Pharmaceuticals, Genomics PLC; and a Life Sciences Industry Consortium of AstraZeneca PLC, Bristol-Myers Squibb Company, GlaxoSmithKline Research and Development Limited, Maze Therapeutics Inc, Merck Sharp & Dohme LLC, Novo Nordisk A/S, Pfizer Inc, Takeda Development Centre Americas Inc.

We thank Social Action for Health, Centre of The Cell, members of our Community Advisory Group, and staff who have recruited and collected data from volunteers. We thank the NIHR National Biosample Centre (UK Biocentre), the Social Genetic & Developmental Psychiatry Centre (King’s College London), Wellcome Sanger Institute, and Broad Institute for sample processing, genotyping, sequencing and variant annotation. This work uses data provided by patients and collected by the NHS as part of their care and support. This research utilised Queen Mary University of London’s Apocrita HPC facility, supported by QMUL Research-IT, http://doi.org/10.5281/zenodo.438045

We thank: Barts Health NHS Trust, NHS Clinical Commissioning Groups (City and Hackney, Waltham Forest, Tower Hamlets, Newham, Redbridge, Havering, Barking and Dagenham), East London NHS Foundation Trust, Bradford Teaching Hospitals NHS Foundation Trust, Public Health England (especially David Wyllie), Discovery Data Service/Endeavour Health Charitable Trust (especially David Stables), Voror Health Technologies Ltd (especially Sophie Don), NHS England (for what was NHS Digital) - for GDPR-compliant data sharing backed by individual written informed consent.

Most of all we thank all of the volunteers participating in Genes & Health.

A favourable ethical opinion for the main Genes & Health research study was granted by NRES Committee London - South East (reference 14/LO/1240) on 16 Sept 2014. Queen Mary University of London is the Sponsor, and Data Controller.

## Data availability

Genes & Health: Individual-level participant data are available to researchers and industry partners worldwide via application to and review by the Genes & Health Executive (https://www.genesandhealth.org/); applications are reviewed monthly. Approved researchers have access to individual-level data in the Genes & Health Trusted Research Environment (TRE) and can request the data files used in this study from the corresponding author(s). All data exports from the Genes & Health TRE are reviewed to prevent release of identifiable individual-level data. Summary data may be exported for cross-cohort meta-analysis or replication and for publication, subject to review

## Competing interests

CM, AD and YJ are current employees of Novo Nordisk Foundation.

---

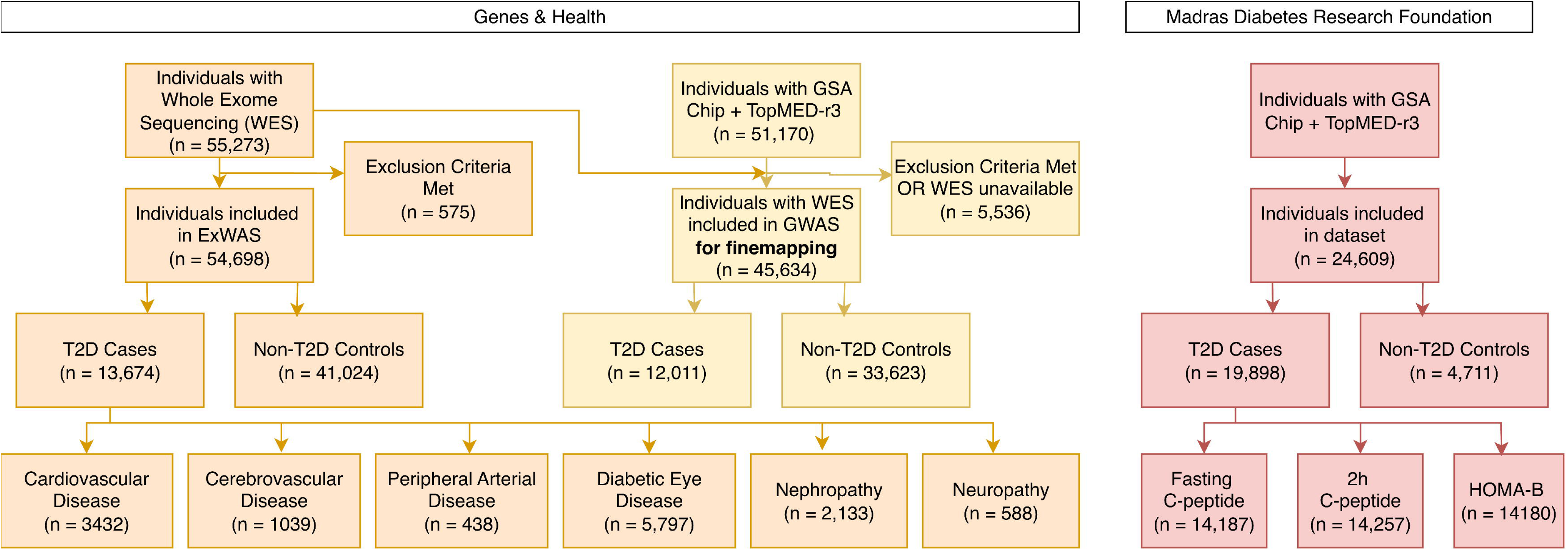

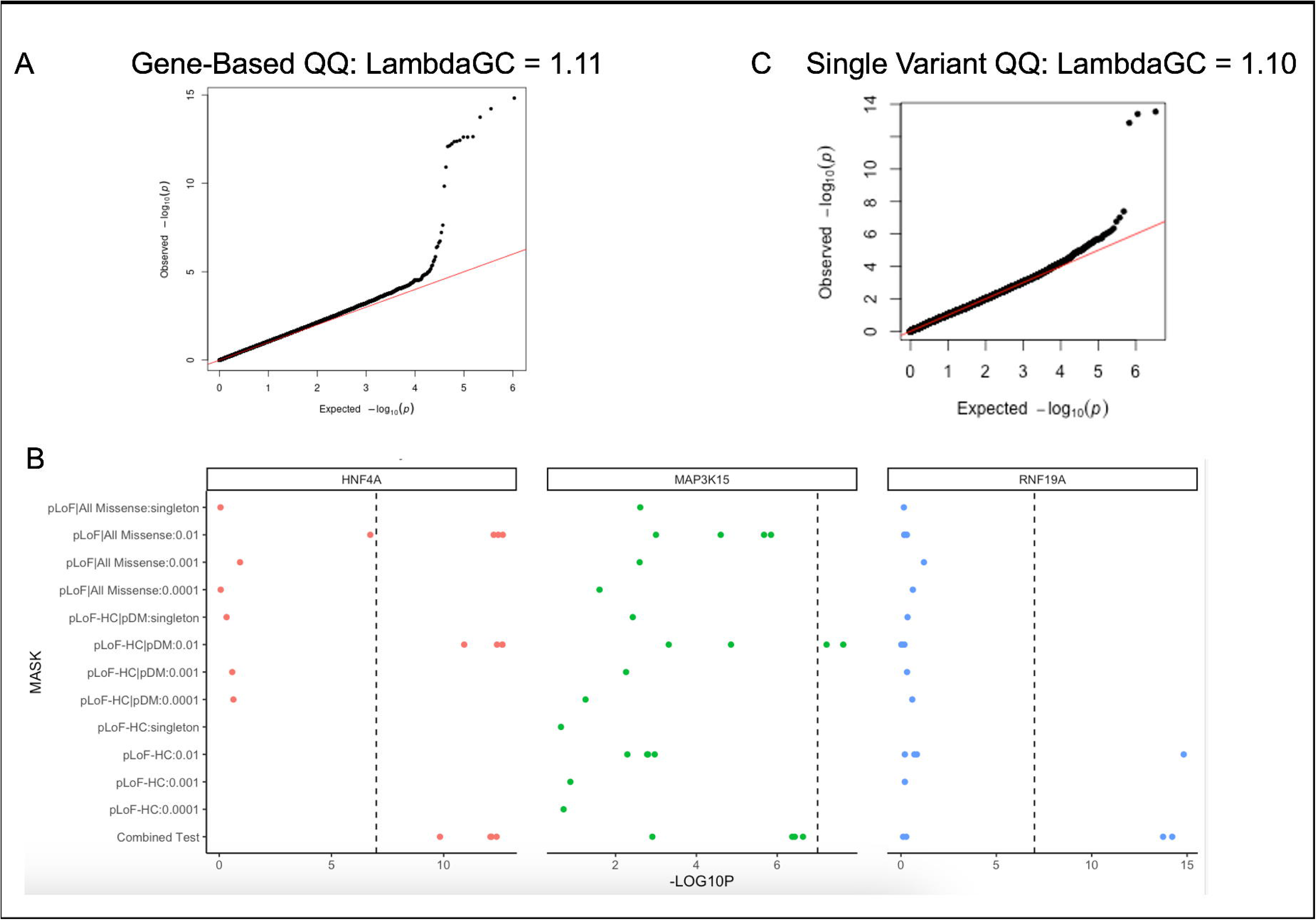

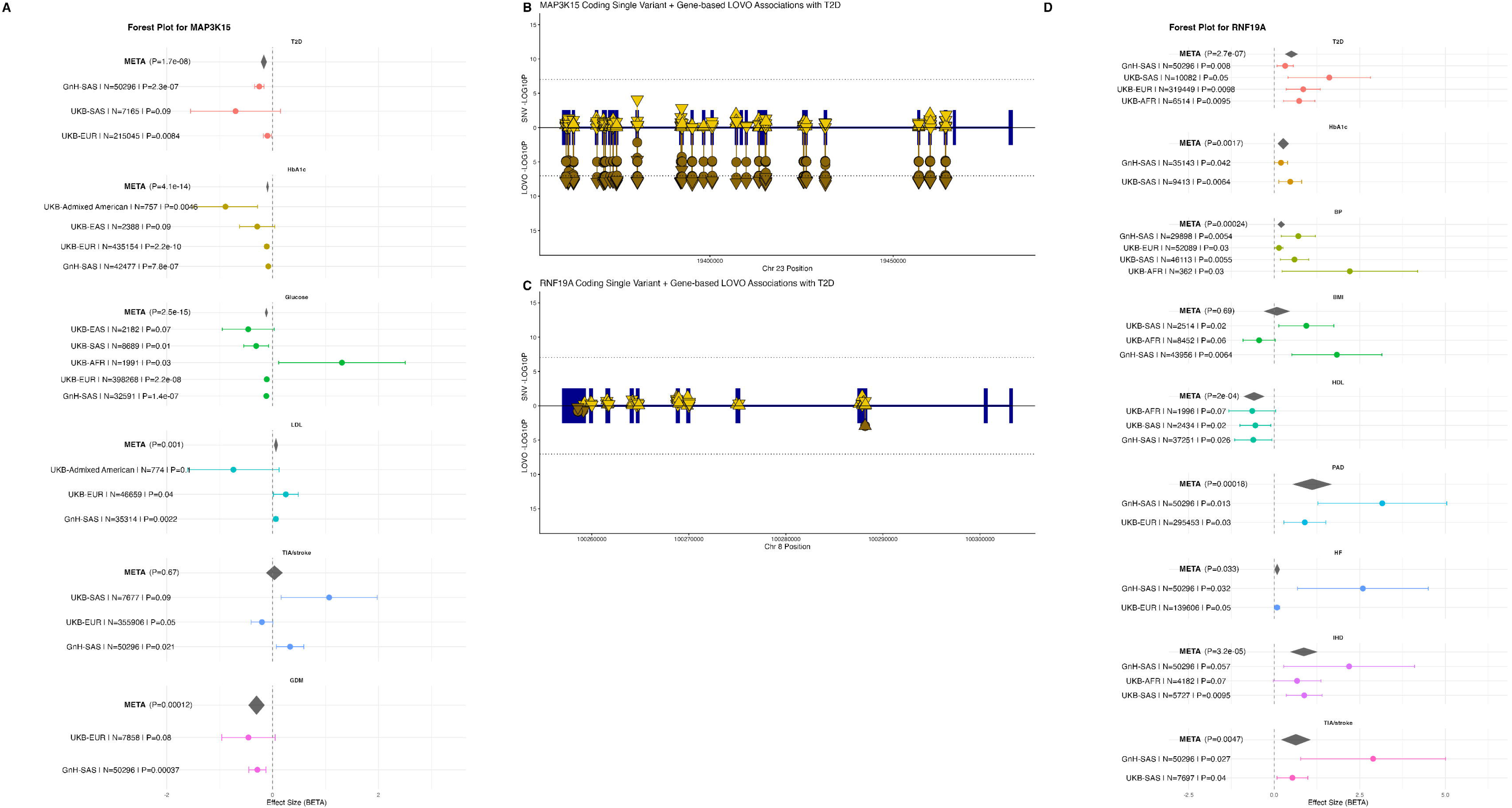

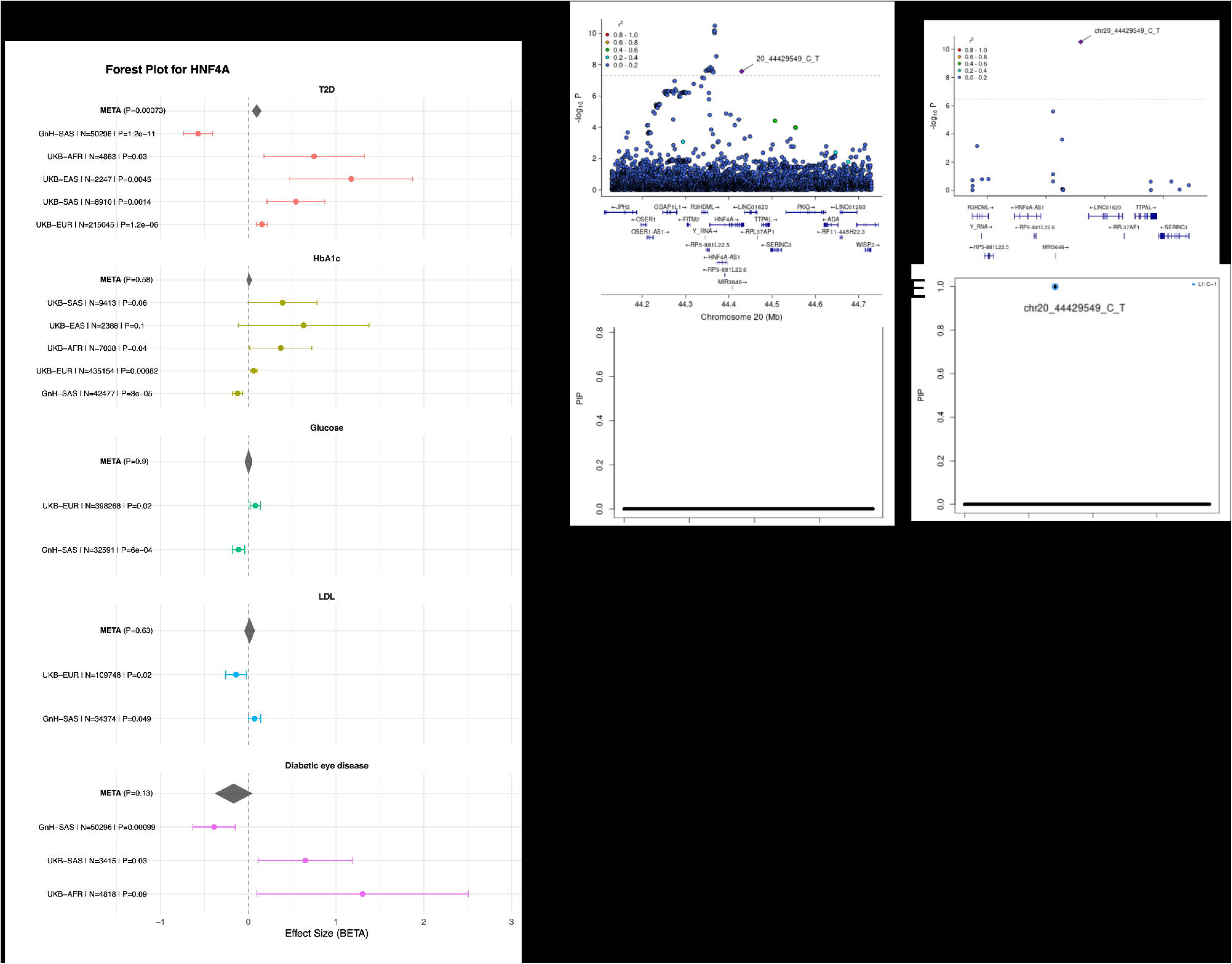

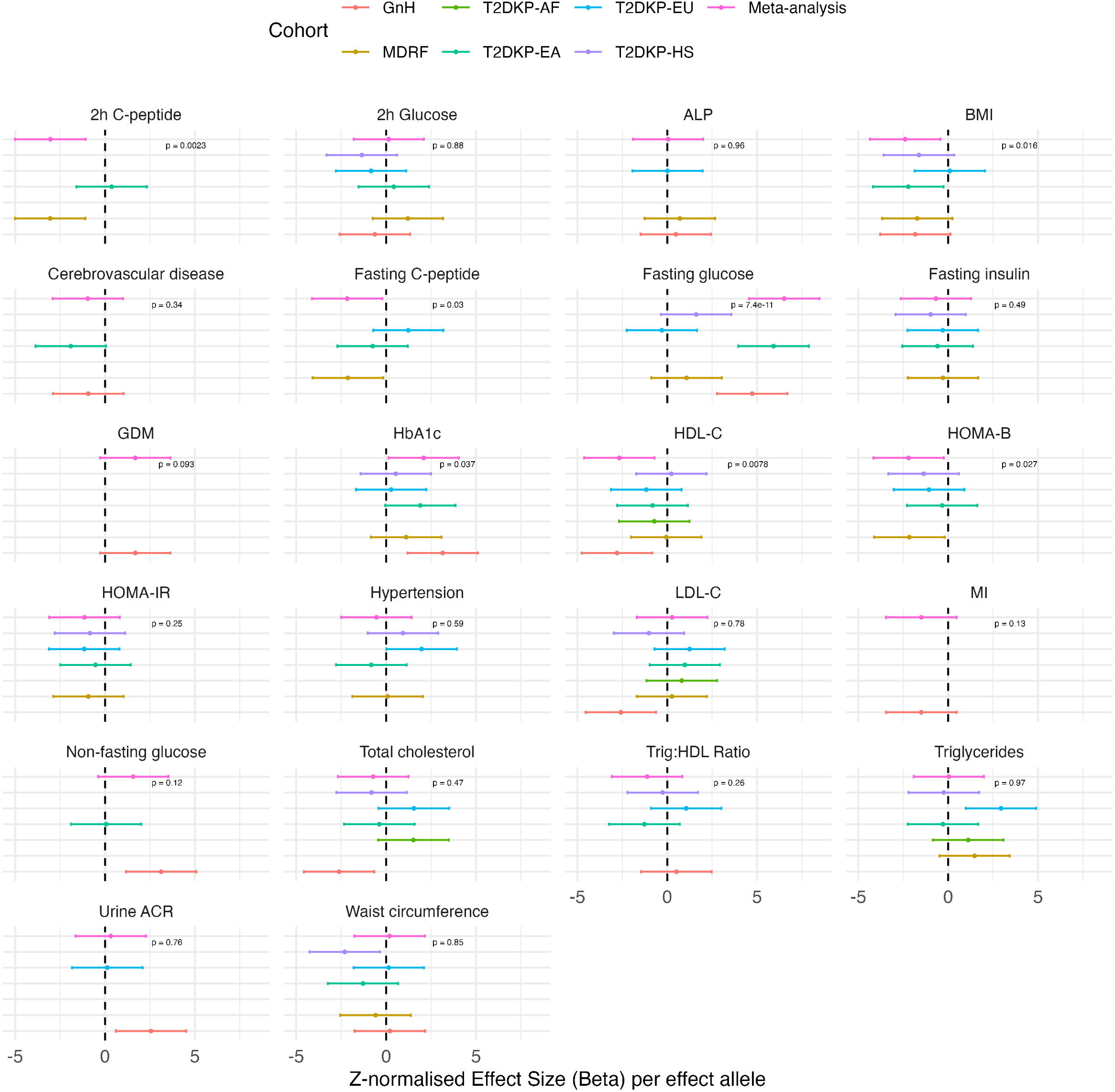

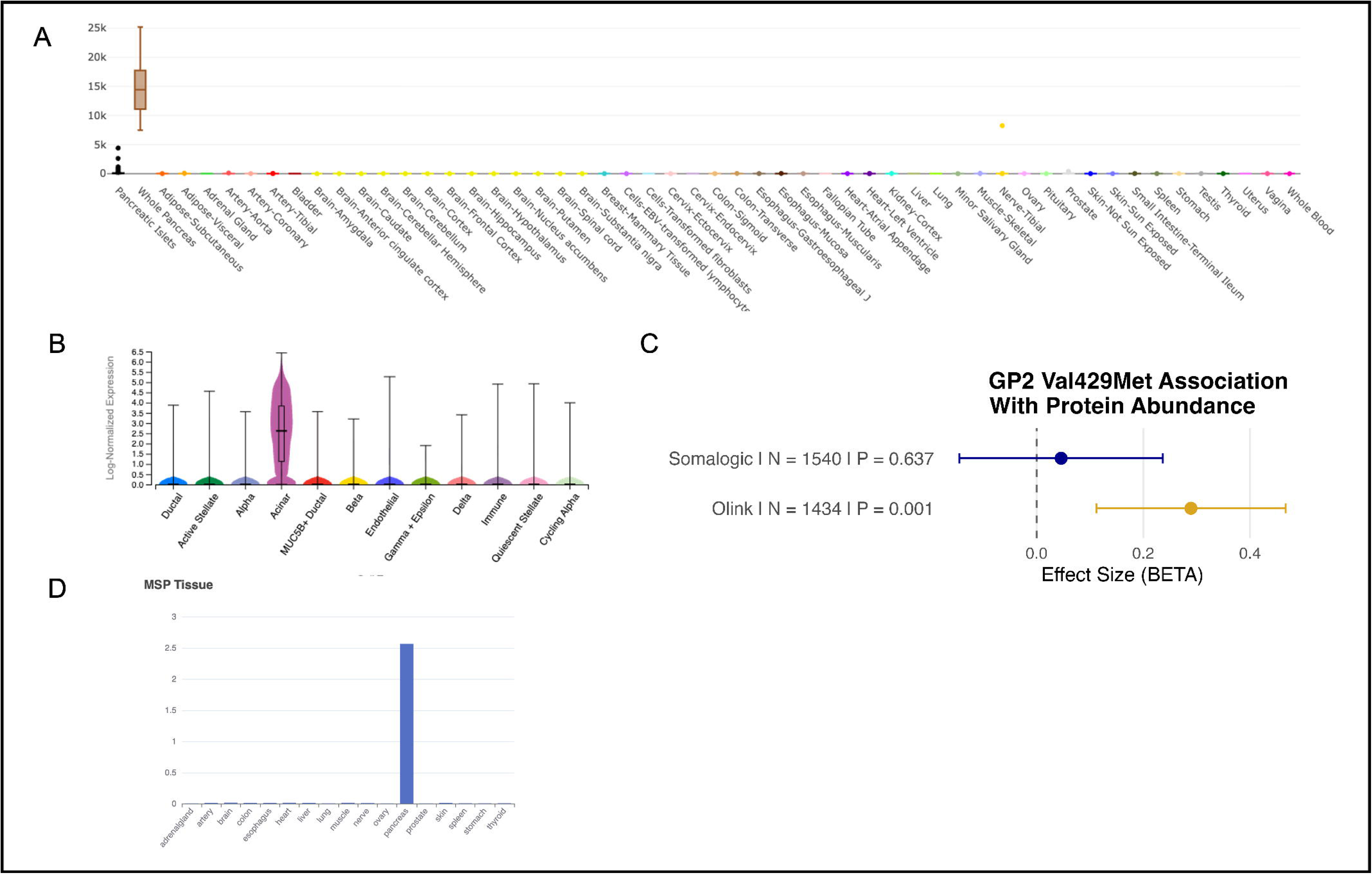

## Notes

### Author Declarations

A favourable ethical opinion for the main Genes & Health research study was granted by NRES Committee London - South East (reference 14/LO/1240) on 16 Sept 2014. Queen Mary University of London is the Sponsor, and Data Controller. The MDRF data used in our study were individual-level data; ie, the analyses were undertaken in a secure cloud environment, including individual-level imputed genotype data linked to clinical metrics (eg HOMA-IR, HbA1c, etc). This data is of course anonymised with no identifiable information available to researchers.

### Summary of Updates

Substantial changes and additional analyses, including: external single variant replicartion in UKBIOBANK for HNF4a P437S external replication of domain-specific HNF4A associations wirh T2D Addition of medication data Addition of multiple external collaborators as part of external replication work

