## Supplementary material for "An ancestry-enriched HNF4A variant and GP2 reveal distinct mechanisms of type 2 diabetes in exome-wide study of 13,674 cases and 41,024 controls"

**Hodgson *et al.***

#### Deleterious, low frequency MAP3K15 variants are associated with lower risk of T2D and its complications

We recapitulate previously reported negative association^1^ between *MAP3K15* variants and type 2 diabetes (**Fig S3A)**. The strongest associations were for the mask capturing deleterious missense variants with MAF < 0.01 (meta-analysed OR = 0.85, 95% CI = 0.80-0.90, p =1.7 x 10^-8^), with no specific variant driving this effect in leave-one-variant-out (LOVO) analysis (**Fig S3B)**. In phenome-wide association testing within Genes & Health meta-analysed with UK Biobank, this mask was significantly associated with lower lifetime median HbA1c (beta = -0.09, 95% CI = -0.12 - -0.07, p = 4.1e-14) lower non-fasting glucose (beta = -0.12, 95% CI = -0.14 - -0.08, p = 2.5 x 10^-15^), and reduced risk of gestational diabetes (OR = 0.74, 95% CI = 0.63 – 0.86, p = 1.2 x 10^-4^).

#### High-confidence loss-of-function RNF19A rare variants drive higher risk of T2D and cardiovascular complications

We identified a novel, exome-wide significant association between HC-pLoF variants with MAF < 0.01 in *RNF19A:pLoF-HC* and type 2 diabetes (GENE_P p value = 1.8x10^-14^ ; however, this does not include an estimate of effect size or direction. Meta-analysed OR (incorporating G&H SKAT test results with direction of effect and multi-ancestry samples from UKBiobank) = 1.65, 95% CI = 1.37 – 2.01, p = 2.8 x 10^-7^). In LOVO analysis, the strongest individual signal arose from a SAS-specific risk-increasing stop-gain mutation (8:100288165:G:A, *RNF19A* Gln4Ter, MAF = 9.1x 10^-6^, beta = 5.36, p = 0.001; MAF in GnOMAD SAS= 3.4 x 10^-5^) resulting in truncation of the protein at the fourth amino acid **(Fig S3C)**.

The effects of this variant could not be meta-analysed in UK-Biobank because no carriers could be identified. However, meta-analysis of *RNF19A* pLoF-HC variants from UKBiobank obtained via the AZPhewas portal^2^ with Genes & Health identified significant associations with increased risk of ischaemic heart disease (OR = 2.38, 95% CI = 1.58 – 3.58, p = 3.2 x 10^-5^) and peripheral arterial disease (OR = 3.04, 95% CI = 1.70 – 5.43, p = 1.8 x 10^-4^), and nominal associations with cerebrovascular disease (OR = 1.89, 95% CI = 1.22 – 2.93, p = 4.7 x 10^-3^) and heart failure (OR = 1.09, 95% CI = 1.01 – 1.18, p = 0.03) (**Fig S3D)**. Significant continuous trait associations including HbA1c (beta = 0.27, 95% CI = 0.10 – 0.43 p = 1.7 x 10^-3^), systolic blood pressure (beta = 0.21, 95% CI = 0.10 – 0.32, p =2.4 x 10^-4^), and HDL-cholesterol (beta = -0.59, 95% CI = -0.90 - -0.28, p = 1.97 x 10^-4^) were also observed after meta-analyses with UKBiobank **(Fig S3D).**

However, the low frequency of most damaging variants, lack of consistency in strength of association across LOVO masks, and nominal significance of metabolic trait associations mean these findings should be viewed as exploratory, with further replication in independent cohorts needed before meaningful conclusions can be drawn.

*HNF4A Pro437Ser Variant Details*

HNF4a Pro437Ser is slightly more common in British Bangladeshis than Pakistanis in Genes & Health (MAF 0.0098 vs 0.0063)**.** Notably it was not included in any UK Biobank gene-burden sets on the AZ Phewas portal^2^ and also not represented in publicly available gene-level testing^3^ since they are from EUR cohorts. This variant is not linked to nearby variants, including those with significant T2D associations (maximum R^2^ = 0.002, **Fig S4B-E).** To identify likely causal variants, we performed fine mapping using SuSIE v0.12.35 on imputed genotype data after performing a matched genome-wide association study. Fine-mapping supported rs150776703 as the likely causal variant at this locus (posterior inclusion probability = 0.997 in imputed data and 1.0 in exome sequencing data; Supplementary Table S6–7).

*HNF4A Domain-specific associations with T2D across cohorts*

In Genes & Health, rare missense variants in the non-repressor domain were associated with increased risk of type 2 diabetes (OR = 1.41, 95% CI = 1.05 – 1.88, p = 0.02**),** consistent with previous observations for *HNF4A*. In contrast, variants within the repressor domain were associated with reduced risk, but this effect was driven by inclusion of Pro437Ser (OR = 0.52, 95% CI = 0.43-0.62, p = 1.6x10^-11^. Exclusion of this variant abolished the protective association (OR = 0.85, 95% CI = 0.53-1.36, p = 0.50), indicating that the effect was not shared across other repressor domain variants. Consistent findings were observed in a meta-analysis between UK Biobank (n= 459,903), and AMP-T2D (Accelerating Medicines Partnership in Type 2 Diabetes, n = 20,074), where repressor domain variants excluding Pro437Ser were associated with increased risk of type 2 diabetes (OR = 1.51, 95%CI = 1.12-2.05, P = 0.008). Together, these results suggest that the protective association is specific to Pro437Ser and cannot be attributed to variant location within the *HNF4A* repressor domain.

*GP2 Allele Frequency Differences across populations are unlikely to be attributable to chance*

Using population genetic simulation methods similar to those used to understand genetic drift in Greenlanders ^27^, we found that under a simple model of neutral evolution, the observed allele frequency differentiation between Europeans and South Asians (p = 0.0012), and between Europeans and East Asians (p = 0.0011), was unlikely to have arisen by chance alone.

*GP2 Tissue expression from public repositories*

Expression quantitative trait data from GTEx^31^, the Translational Human Pancreatic Islet Genotype Tissue-Expression Resource (TIGER)^32^, and Pankbase^33^ indicateds*GP2* expression is largely restricted to the pancreas, with minimal expression in islet cells, and predominant expression in acinar cells (**Figure S6A/B).**

*Sex-stratified analyses*

We observed no sex-specific effects in *HNF4A* metabolic trait associations, nor its association with T2D (**Table S4).**

*Sensitivity analysis: association with autozygosity*

We explored the association of identified variants of interest with the autozygosity marker FROH (fraction of genome in runs of homozygosity), a measure of the proportion of the genome which is in long runs of homozygosity mediated via identity-by-descent. Since markedly different rates of autozygosity are observed in different ancestral groups, we stratified results by ancestry as well as sex, in addition to pooled analyses. We used pre-called FROH metrics as defined in^3^ and examined the association of FROH with variants of interest. Neither HNF4A Pro437Ser nor GP2 Val429Met were associated with FROH in either the whole sample, nor subsamples stratified by sex or ancestry (max p_association across all 9 possible sex*ancestry combinations = 0.12).

*Sensitivity analyses for glycaemia*

We explored whether two variants characterised in detail in this analysis were associated with metrics of glycaemia in non-diabetic participants, defined according to MAGIC consortium criteria described elsewhere^4^. In brief, we restricted HbA1cs included in analysis to 33,890 individuals with HbA1c values which pre-dated either earliest diagnosis of T2D or earliest date of medication, after excluding values in pregnancy windows. We observed consistency in results for both *HNF4A* pro437Ser (beta = -1.4 mmol/mol, 95% CI = -2.3 - -0.75, p = 0.001) and *GP2* Val429Met (beta = 0.45 mmol/mol, 95% CI = 0.007 – 0.91, p = 0.04).
